# Diurnal Glycemic Redistribution in Obesity Despite Matched Mean Glucose

**DOI:** 10.64898/2026.10.27.26366601

**Authors:** Ren Zhang

## Abstract

**Background and Aims:** Mean glucose and HbA1c reflect average glycemic exposure but do not capture its 24-hour distribution. When average glycemia is comparable, whether obesity is associated with a distinct temporal glucose pattern remains unclear. We compared continuous glucose monitoring (CGM) profiles in lean adults and adults with obesity matched for mean glucose.

**Methods:** In a cross-sectional AI-READI analysis, lean adults (BMI 18.5–<25 kg/m²) were matched one-to-one to adults with obesity (BMI ≥30 kg/m²) by recruitment center and clinical group, with calipers for age, mean glucose and glucose standard deviation. CGM profiles were compared by clock time and relative to wearable-defined sleep onset. Sensitivity analyses tightened mean-glucose calipers from 10 to 2 mg/dL.

**Results:** Among 261 matched pairs, mean glucose (lean versus obesity: 122.34 versus 122.82 mg/dL) and HbA1c (5.73% versus 5.78%) were comparable. Obesity was associated with higher nighttime glucose (00:00–06:00; +6.18 mg/dL) and lower afternoon glucose (12:00–18:00; −5.11 mg/dL; both adjusted P<0.001). Sleep-aligned analyses supported the nocturnal pattern, including among healthy participants and those with prediabetes. These differences persisted with a 2 mg/dL mean-glucose caliper (nighttime +3.86 mg/dL; afternoon −4.27 mg/dL; both adjusted P<0.001).

**Conclusion:** Despite comparable mean glucose and HbA1c, obesity is associated with diurnal glycemic redistribution, characterized by higher nocturnal and lower daytime glucose. These findings support examining glucose timing alongside conventional measures of average glycemia.

## Introduction

Diabetes and prediabetes are major public health challenges, with diabetes prevalence increasing worldwide and a substantial burden of both conditions in the United States [1, 2]. Understanding glucose dysregulation requires characterization beyond conventional measures of average exposure. Although HbA1c and mean glucose summarize overall glycemia, they do not describe how glucose is distributed across the day [3]. Continuous glucose monitoring (CGM) enables examination of glucose profiles under free-living conditions, providing information that aggregate glycemic measures may obscure [4–6]. Glucose regulation varies across the 24-h cycle through the combined influences of circadian physiology, feeding, and sleep [7, 8]. Experimental studies have documented obesity-associated alterations in temporal glucose regulation [9, 10], while observational CGM studies have characterized glucose profiles in relation to adiposity and glycemic status [11–13]. These complementary observations provide a rationale for examining not only the magnitude of glucose exposure but also its temporal organization in obesity.

Among individuals with comparable average glycemia, it remains unclear whether obesity is associated with a distinct temporal distribution of glucose. Comparisons that do not account for average glycemia may conflate higher overall glucose with differences in its temporal pattern. The question is therefore not simply whether glucose is higher in obesity, but whether comparable average exposure conceals a different distribution across the day. A related question is whether such differences are detectable even among individuals with normoglycemia. A glycemia-matched comparison within clinical categories can address these questions, while individual sleep measurements provide a complementary temporal reference beyond clock time.

Here, we used linked clinical, CGM, and wearable-derived sleep data from AI-READI to compare lean participants and participants with obesity matched on mean glucose within clinical categories [14, 15]. We examined clock-time profiles, glucose trajectories aligned to sleep onset, and glucose concentrations during detected sleep, and assessed robustness to progressively tighter mean-glucose matching. The study addressed whether obesity is associated with diurnal glucose redistribution despite comparable average glycemic exposure.

## Materials and Methods

### Data source and study design

This cross-sectional secondary analysis used linked clinical measurements, continuous glucose monitoring (CGM), and wearable-derived sleep data from AI-READI (version 3.0.0). Recruitment and data collection have been described previously [14, 15]. Clinical categories were retained from the source dataset. The prediabetes/lifestyle-controlled category includes both prediabetes and lifestyle-controlled diabetes; the healthy category is reported as a source classification. Participants with BMI 18.5 to <25 kg/m² were matched one-to-one without replacement to those with BMI ≥30 kg/m². Eligibility required linked CGM and sleep records, available matching variables, and complete clock-hour profiles as defined below. Matching was exact by recruitment center and clinical category, with maximum within-pair differences of 10 years for age, 10 mg/dL for mean glucose, 5 mg/dL for glucose standard deviation, and 1 eligible monitoring day. Within each center–category stratum, minimum-cost assignment selected pairs using summed squared differences in age, mean glucose, glucose standard deviation, and eligible-day count, each standardized by its standard deviation in the eligible participant pool. The single matched pair in the source insulin-dependent category was excluded. HbA1c and sex were not matching variables. The present study was approved by the Institutional Review Board of Wayne State University.

### CGM and wearable data processing

CGM and wearable records were aligned by participant and timestamp. Data were collected using Dexcom G6 and Garmin vívosmart 5 devices during a planned 10-day monitoring period, with CGM sampled every 5 min. Timestamps were harmonized in UTC and converted to recruitment-center local time, accounting for daylight-saving transitions. Records with missing or invalid timestamps or glucose values were excluded, duplicate participant–timestamp records were removed, and glucose values of 40–401 mg/dL were retained. Missing readings were not interpolated.

Garmin records were used to identify detected sleep and main sleep episodes. Light, deep, and REM intervals defined detected sleep. Invalid and duplicate intervals were removed, and participants with remaining overlapping intervals were excluded. Consecutive sleep-stage or awake intervals separated by ≤1 h were grouped into episodes, bounded by the first and last detected sleep intervals. Eligible main episodes spanned 3–12 h, contained ≥2 h of detected sleep, and fell within a local noon-to-noon window. When multiple episodes qualified, the episode containing the longest detected sleep duration was selected. Monitoring-day eligibility required adequate CGM coverage during and outside sleep. Each noon-to-noon window required ≥70% of expected readings both overall and during detected sleep, together with ≥24 detected-sleep readings and ≥72 outside-sleep readings. Expected counts were calculated from interval durations and the 5-min sampling frequency. Participants required at least three eligible days.

### Glucose profile and sensitivity analyses

Participant-level glycemia was calculated by averaging daily glucose means and daily SDs across all eligible noon-to-noon windows, with equal weighting of days; these values were used for matching. For daily comparisons, windows were indexed from each participant’s first eligible day without compressing missing calendar days. Comparisons across the first 10 indexed windows included only pairs with data for both members. Clock-hour profiles were constructed by averaging daily hourly means within participants, requiring ≥8 readings per hourly bin and ≥3 contributing days for every clock hour. Mean glucose for nighttime (00:00–06:00), morning (06:00–12:00), afternoon (12:00–18:00), evening (18:00–24:00), and remaining hours (06:00– 24:00) was calculated by equally weighting the corresponding hourly means. Interval starts were inclusive and ends exclusive. The night-minus-afternoon contrast was calculated within participants. Clock-time analyses included the full cohort and each retained clinical category.

Sleep-related analyses evaluated onset-aligned profiles and detected-sleep averages. Profiles used 1-h bins from 3 h before to 6 h after the first detected sleep interval of the main episode, requiring ≥8 readings per bin on ≥3 days for every bin in both members of a pair. Post-onset bins had to end within the episode but could include awakenings. Daily mean glucose during detected sleep, including naps, and outside detected sleep was averaged across eligible days. Outside-sleep readings excluded the main episode and all Garmin-staged intervals and were not considered confirmed wakefulness. Sensitivity analyses repeated matching from the eligible participant pool at mean-glucose calipers of 10, 8, 6, 5, 4, and 2 mg/dL, retaining all other criteria and standardization scales. Outcomes were nighttime glucose, afternoon glucose, and the night-minus-afternoon contrast; matched samples overlapped across thresholds.

### Statistical analysis

Statistical comparisons used participant-level summaries while preserving matched pairs. Group means are presented with SEM. Between-group differences, calculated as obesity minus lean, were evaluated using two-sided paired *t*-tests, with unadjusted 95% confidence intervals reported for paired mean differences. Repeated days and individual CGM readings were not treated as independent observations. Multiplicity was addressed using Holm correction within each analysis family: 10 daily mean-glucose comparisons, 96 clock-hour comparisons, 16 clock-interval comparisons, 36 sleep-onset comparisons, and five clock-defined and wearable-defined exposure comparisons. Sensitivity analyses applied Holm correction across the six matching thresholds separately for each outcome. A *P* value <0.05 was considered statistically significant. Processing, analysis, and visualization were performed in Python using NumPy, pandas, SciPy, statsmodels, and Matplotlib.

## Results

### A glycemia-matched cohort of lean participants and participants with obesity

Average glycemic measures summarize glucose exposure but do not resolve its temporal pattern. To examine glucose profiles at comparable levels of overall glycemia, we constructed a matched cohort from AI-READI (Fig. 1A). Lean participants (18.5 ≤ BMI <25 kg/m²) were matched one-to-one with participants with obesity (BMI ≥30 kg/m²), exactly by recruitment center and clinical category. Matching permitted a maximum within-pair difference of 10 mg/dL in mean continuous glucose monitoring (CGM) glucose, with additional constraints on age, glucose standard deviation, and eligible monitoring days. After exclusion of one insulin-dependent pair, the final cohort comprised 261 matched pairs (522 participants): 124 in the healthy category, 82 in the prediabetes/lifestyle-controlled category, and 55 in the non-insulin medication category.

**Figure 1.**
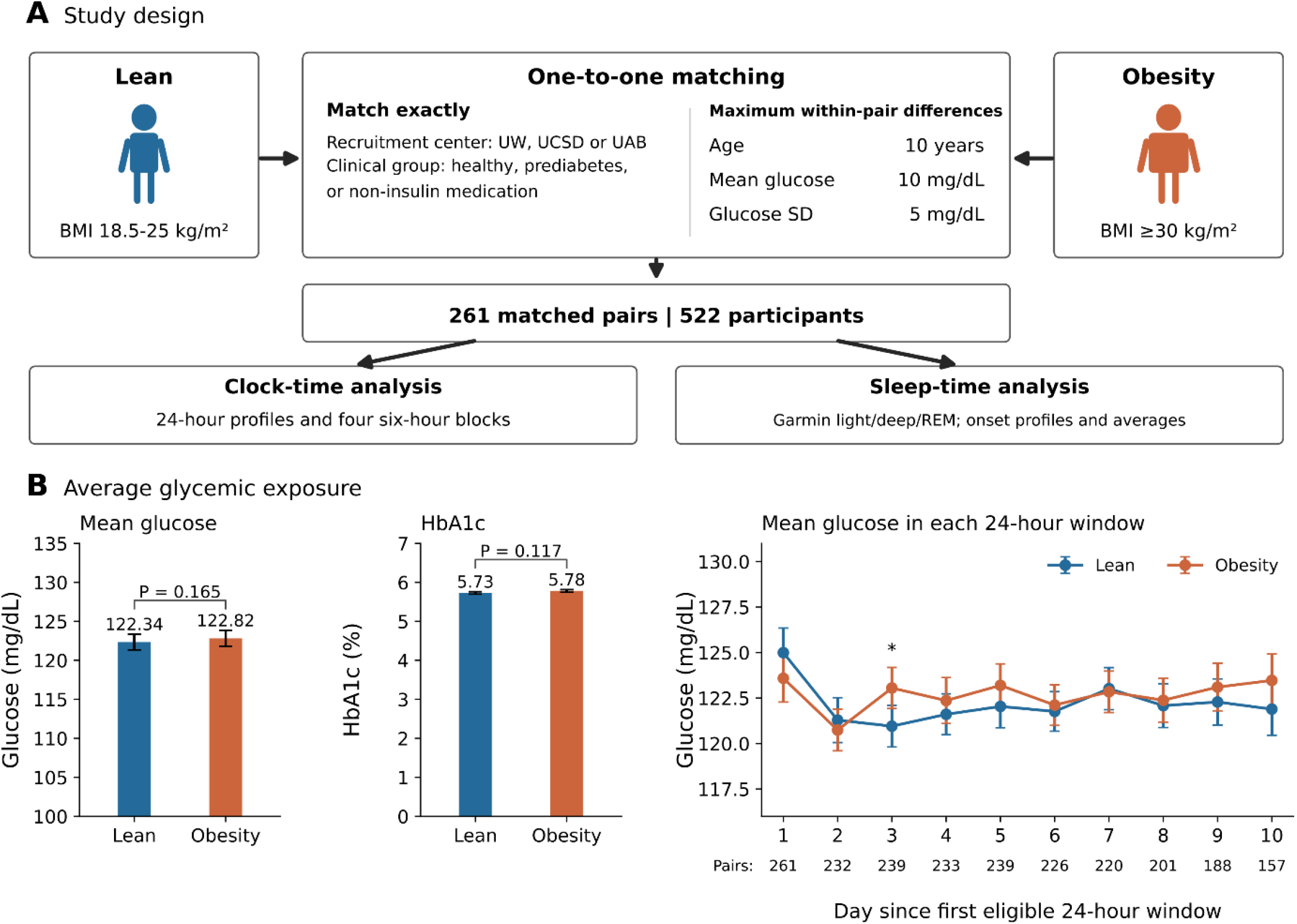
Study design and average glycemic exposure. **(A)** Lean participants (18.5 ≤ BMI <25 kg/m²) and participants with obesity (BMI ≥30 kg/m²) were matched by recruitment center and clinical category, yielding 261 pairs (522 participants). Maximum within-pair differences were 10 years for age, 10 mg/dL for mean glucose, 5 mg/dL for glucose SD, and 1 eligible monitoring day. Analyses used clock time and Garmin-derived sleep timing. **(B)** Overall mean CGM glucose, HbA1c, and daily mean glucose in noon-to-noon windows, indexed from the first eligible window without compressing missing days. Values are mean ± SEM; numbers below the daily plot indicate contributing pairs. Overall paired P values are unadjusted; daily comparisons were Holm-adjusted across 10 days (*adjusted P <0.05). HbA1c was not matched. Blue, lean; orange, obesity.

Overall mean CGM glucose was comparable between lean participants and participants with obesity (122.34 versus 122.82 mg/dL, respectively; *P* = 0.165; Fig. 1B). HbA1c, which was not used for matching, averaged 5.73% and 5.78%, respectively (*P* = 0.117). Across the 10 indexed noon-to-noon monitoring windows, only the third window showed a statistically significant between-group difference after Holm correction, with higher mean glucose in participants with obesity (adjusted *P* <0.05). The number of contributing matched pairs ranged from 157 to 261 across these windows (Fig. 1B). This matched cohort therefore provided a basis for examining obesity-associated differences in glucose profiles at comparable levels of overall glycemia.

### Obesity is associated with diurnal glucose redistribution despite comparable mean glycemia

To determine whether comparable average glycemia concealed differences in glucose distribution across the day, we examined CGM profiles by local clock hour. In the full matched cohort, the profiles diverged in a time-dependent manner despite comparable overall mean glucose (Fig. 2A). Glucose concentrations were higher in participants with obesity during clock hours 00:00–06:00 (all adjusted *P* <0.001). The profiles converged during the morning, followed by a pronounced early-afternoon rise in lean participants that was less evident in participants with obesity. Consequently, the direction of the difference reversed at 12:00–15:00, when glucose was lower in participants with obesity (all adjusted *P* <0.001). Glucose increased in both groups during the evening, with concentrations again higher in participants with obesity at 22:00 and 23:00 (both adjusted *P* <0.01). Hourly comparisons were Holm-adjusted across the 96 tests shown in Figure 2.

**Figure 2.**
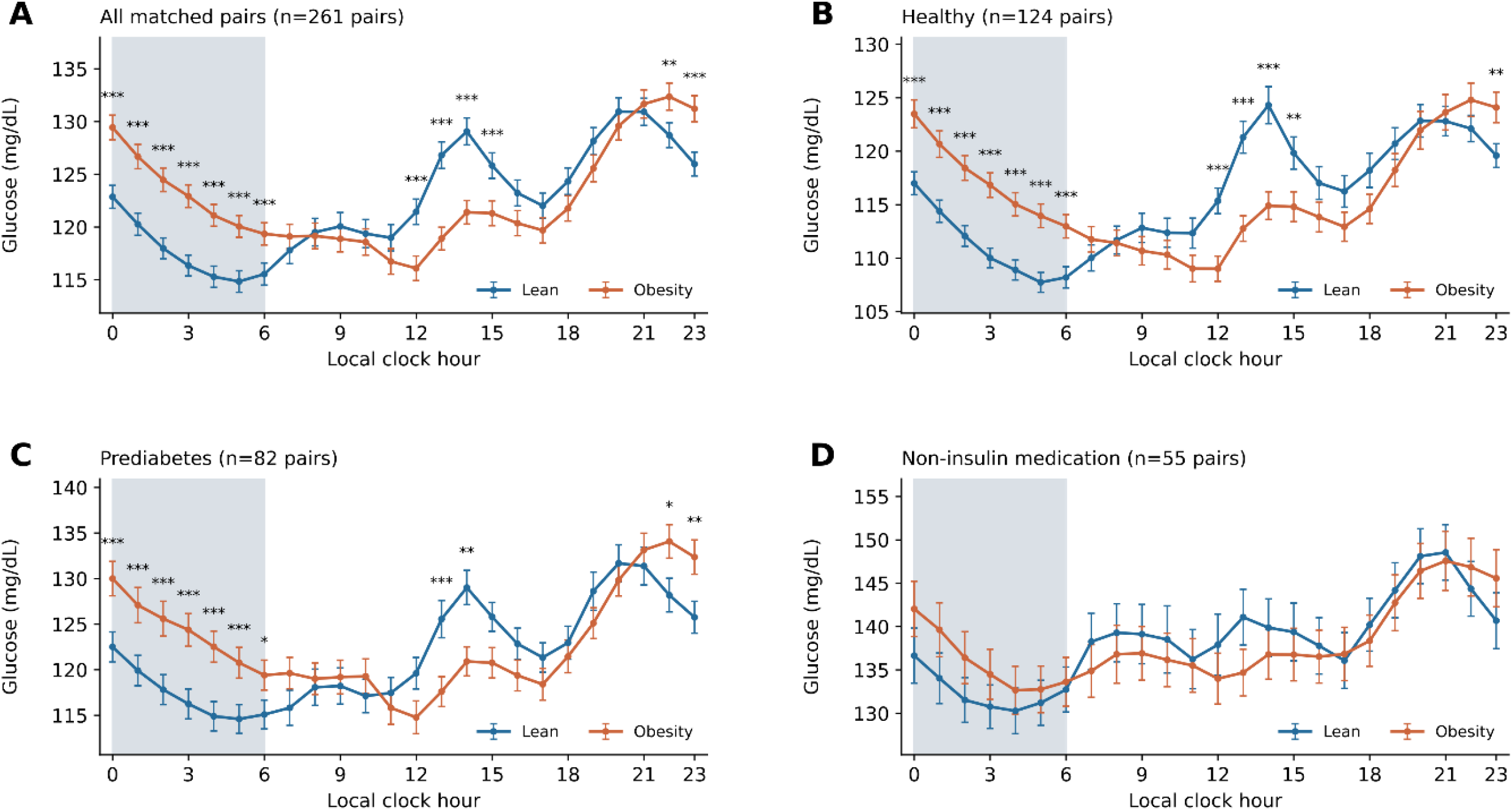
Clock-hour glucose profiles in matched lean and obesity groups. Profiles are shown for **(A)** all matched pairs (n = 261), **(B)** healthy participants (n = 124 pairs), **(C)** prediabetes (n = 82 pairs), and **(D)** participants receiving non-insulin medication (n = 55 pairs). Prediabetes denotes the study’s prediabetes/lifestyle-controlled category. Points and error bars represent mean ± SEM; shading indicates 00:00–06:00. Hourly between-group comparisons used paired t-tests with Holm adjustment across 96 comparisons. *Adjusted P <0.05; **P <0.01; ***P <0.001. Blue, lean; orange, obesity.

The overnight elevation and early-afternoon reduction were also evident in the healthy and prediabetes/lifestyle-controlled categories, comprising 124 and 82 matched pairs, respectively (Fig. 2B,C). Both categories showed significant between-group differences during these periods, together with higher late-evening glucose in participants with obesity. In contrast, none of the hourly comparisons reached statistical significance after multiplicity correction in the 55 pairs receiving non-insulin medication (Fig. 2D). These findings identify an obesity-associated redistribution of glucose across the day, rather than a uniform elevation, within a cohort matched on average glycemia.

### Higher nighttime but lower afternoon glucose in obesity

To quantify the clock-hour differences over broader periods of the day, we summarized glucose profiles across four consecutive 6-h intervals: nighttime (00:00–06:00), morning (06:00–12:00), afternoon (12:00–18:00), and evening (18:00–24:00). Interval-specific comparisons were Holm-adjusted across 16 tests. In the full matched cohort, mean glucose was 6.18 mg/dL higher at night and 5.11 mg/dL lower in the afternoon in participants with obesity than in lean participants (both adjusted *P* <0.001; Fig. 3A). Morning and evening glucose did not differ significantly between groups. The corresponding between-group difference in the night-minus-afternoon contrast was 11.29 mg/dL, quantifying the opposing differences across these periods (Fig. 6, primary analysis).

**Figure 3.**
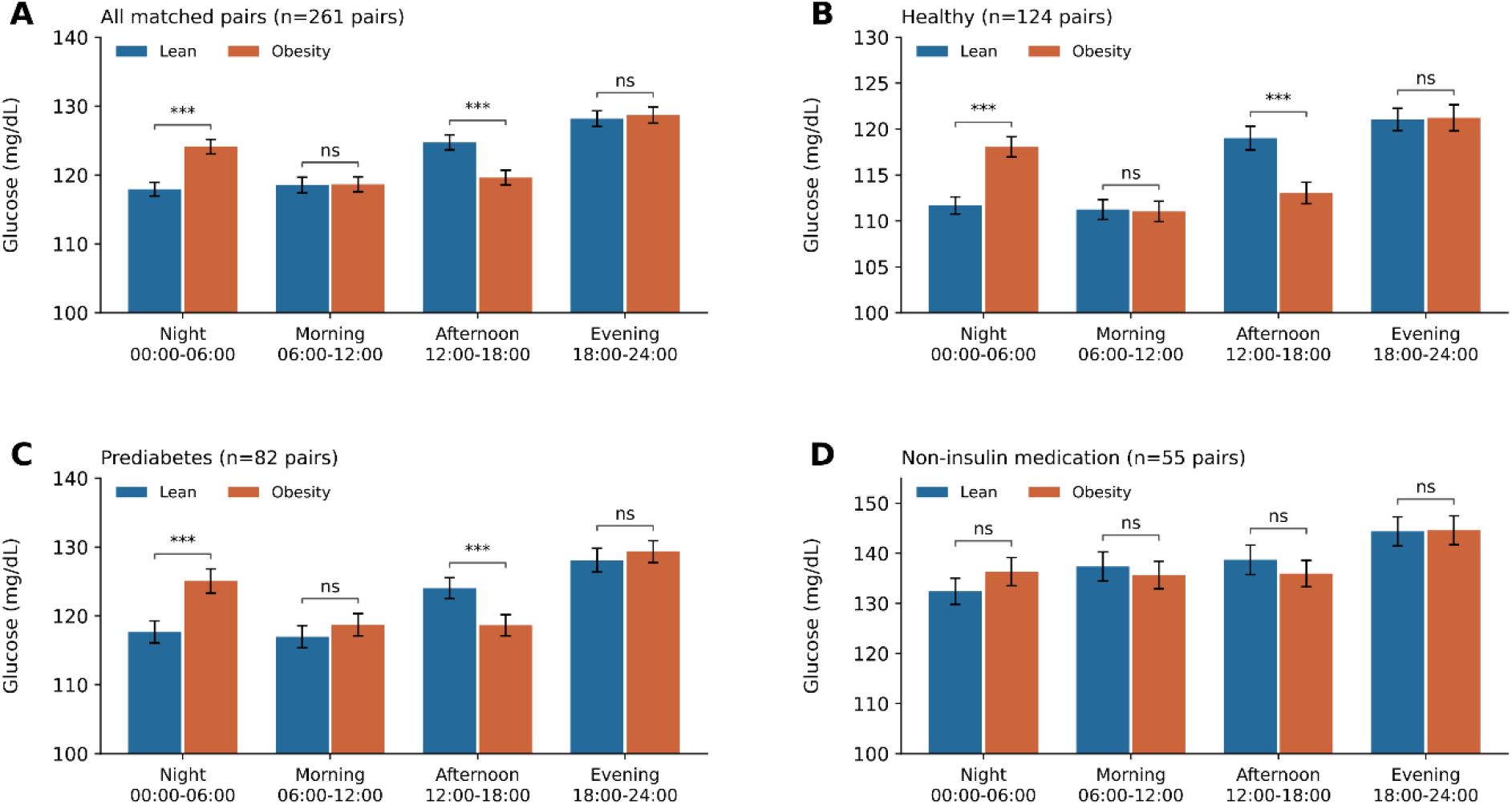
Glucose levels across four clock-time intervals in matched lean and obesity groups. Mean glucose during four 6-h intervals is shown for **(A)** all matched pairs (n = 261), **(B)** healthy participants (n = 124 pairs), **(C)** prediabetes (n = 82 pairs), and **(D)** participants receiving non-insulin medication (n = 55 pairs). Prediabetes denotes the study’s prediabetes/lifestyle-controlled category. Bars represent mean ± SEM of participant-level averages within each interval. Between-group comparisons used paired t-tests with Holm adjustment across 16 comparisons. ***Adjusted P <0.001; ns, not significant. Glucose axes begin at 100 mg/dL. Blue, lean; orange, obesity.

This nighttime–afternoon pattern was also observed in the healthy and prediabetes/lifestyle-controlled categories: participants with obesity had higher nighttime and lower afternoon glucose than their matched lean counterparts (all adjusted *P* <0.001; Fig. 3B,C). Morning and evening differences were not statistically significant in either category. Among participants receiving non-insulin medication, none of the four interval-specific comparisons reached statistical significance (Fig. 3D). Together, these analyses identify higher nighttime and lower afternoon glucose as the defining features of the obesity-associated pattern at comparable mean glycemia.

### Wearable-derived sleep-onset alignment reveals persistent differences in glucose trajectories

Clock-hour analyses do not account for individual differences in sleep timing. To examine glucose trajectories relative to sleep onset, we aligned CGM profiles to Garmin-derived sleep onset in 250 matched pairs with complete profiles from 3 h before to 6 h after onset (Fig. 4). In the full cohort, glucose was already higher in participants with obesity during the hour preceding sleep onset (adjusted *P* <0.001; Fig. 4A). The obesity-group profile reached its highest level within the analyzed window during the first post-onset hour. Although glucose subsequently declined in both groups, concentrations remained higher in participants with obesity in each of the six post-onset hourly intervals (all adjusted *P* <0.001). Comparisons were Holm-adjusted across 36 tests.

**Figure 4.**
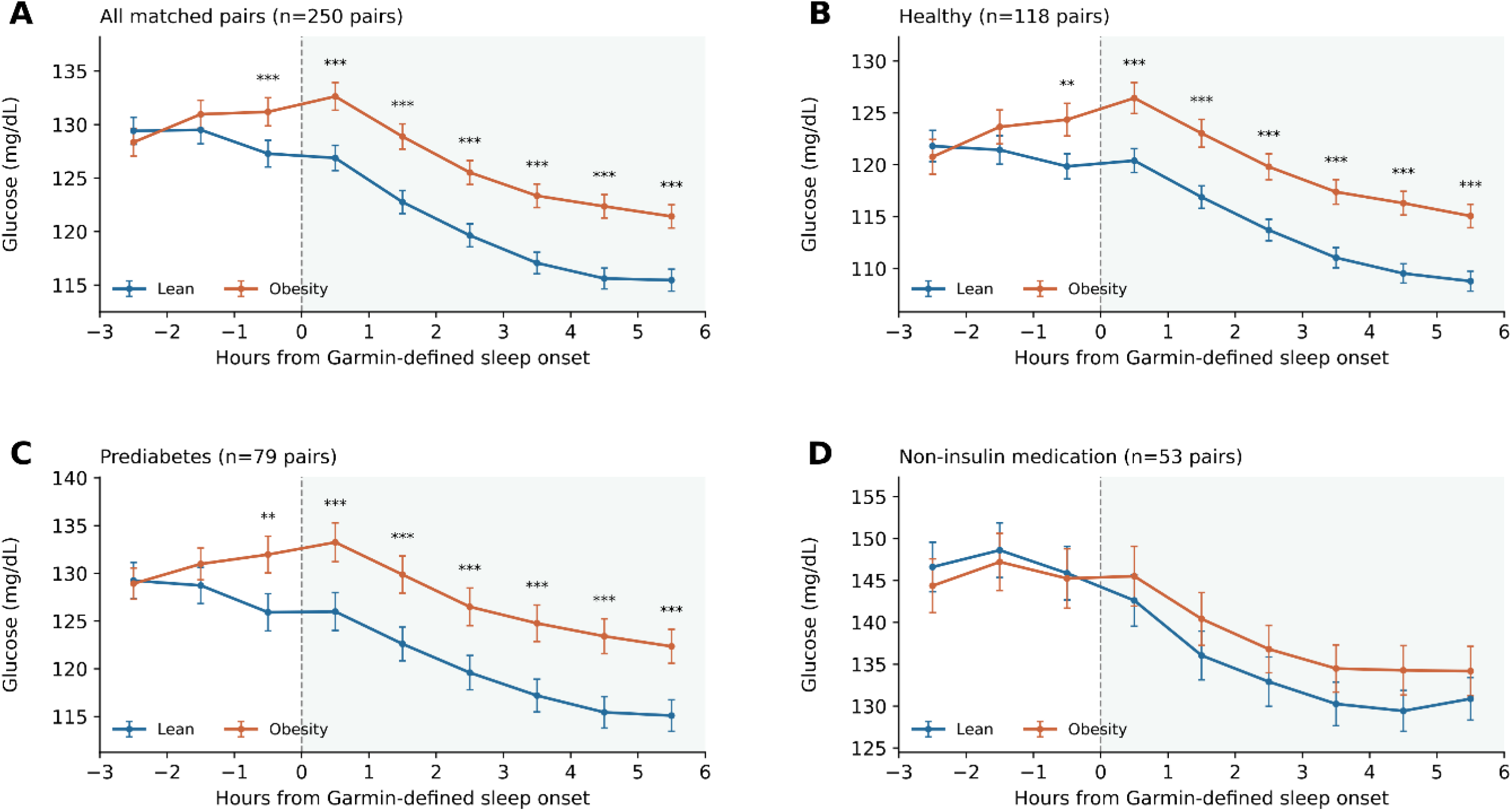
Glucose profiles aligned to Garmin-defined sleep onset. Profiles are shown for **(A)** all matched pairs (n = 250), **(B)** healthy participants (n = 118 pairs), **(C)** prediabetes (n = 79 pairs), and **(D)** participants receiving non-insulin medication (n = 53 pairs). Points represent mean ± SEM in 1-h bins, with ≥3 contributing days per participant per bin. Dashed lines mark sleep onset; shading denotes post-onset intervals, which may include awakenings. Between-group comparisons used paired t-tests with Holm adjustment across 36 comparisons. **Adjusted P <0.01; ***P <0.001. Prediabetes denotes the study’s prediabetes/lifestyle-controlled category. Blue, lean; orange, obesity.

The same pattern was evident in the healthy and prediabetes/lifestyle-controlled categories, comprising 118 and 79 matched pairs, respectively (Fig. 4B,C). In both categories, glucose was higher in participants with obesity during the hour preceding sleep onset (both adjusted *P* <0.01) and throughout the six post-onset intervals (all adjusted *P* <0.001). None of the hourly comparisons reached statistical significance after multiplicity correction in the 53 pairs receiving non-insulin medication (Fig. 4D). These post-onset intervals could include awakenings and therefore did not represent uninterrupted sleep. Together, these findings extend the clock-time results by demonstrating that the glucose differences persisted when profiles were aligned to individual sleep onset.

### Clock-defined night and wearable-detected sleep show concordant glucose differences

Sleep-onset alignment describes glucose trajectories relative to sleep timing but does not isolate intervals of detected sleep. To examine whether the nighttime pattern extended to wearable-detected sleep, we compared mean glucose across clock-defined and Garmin-derived intervals in all 261 matched pairs (Fig. 5). Despite comparable daily mean glucose, participants with obesity had higher glucose during clock-defined night (00:00–06:00) and lower glucose during the remaining hours (06:00–24:00; both adjusted *P* <0.001; Fig. 5A). The wearable-based analysis showed a concordant pattern. Mean glucose was higher in participants with obesity during Garmin-detected sleep, comprising light, deep, and REM sleep and including naps, and lower outside detected sleep (both adjusted *P* <0.001; Fig. 5B). Comparisons were Holm-adjusted across the five tests. Intervals outside detected sleep did not necessarily represent confirmed wakefulness. Together, these analyses show that the glucose redistribution remained evident when periods were defined by detected sleep rather than clock time alone.

**Figure 5.**
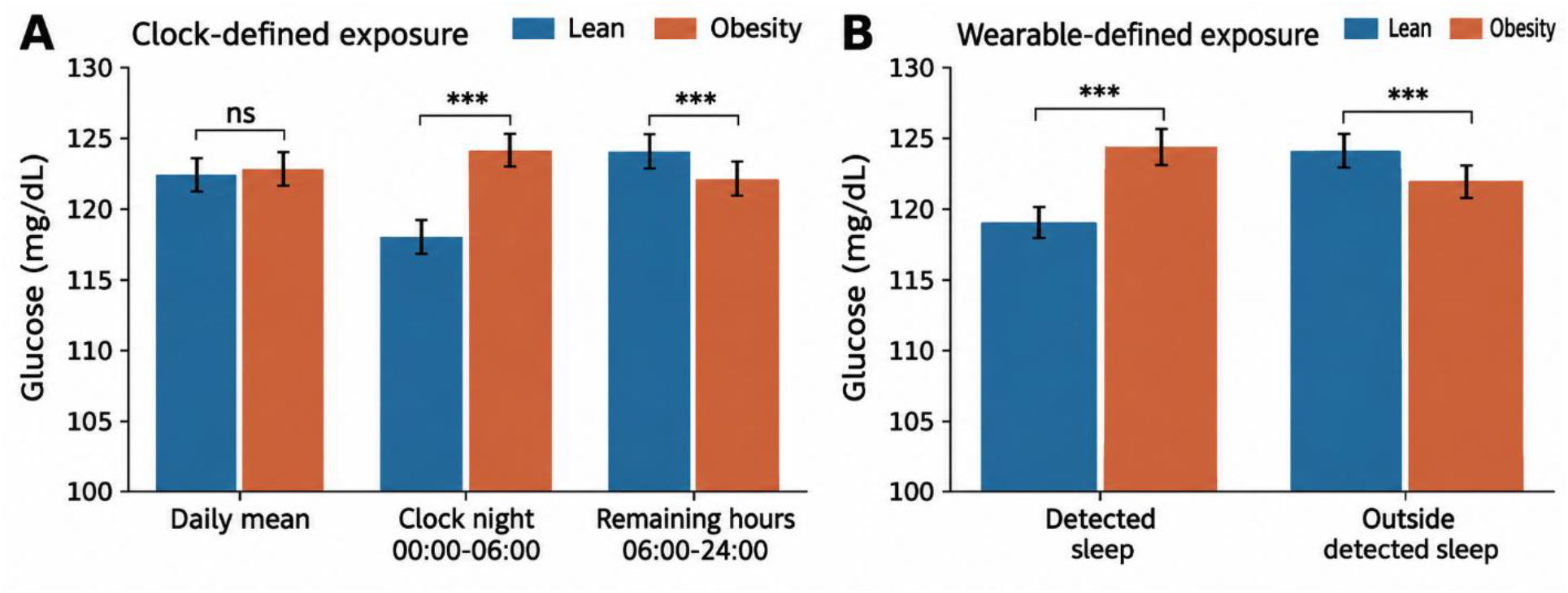
Mean glucose during clock-defined night and Garmin-detected sleep. **(A)** Daily mean glucose and mean glucose during nighttime (00:00–06:00) and remaining hours (06:00–24:00). **(B)** Mean glucose during Garmin-detected sleep (light, deep, and REM sleep, including naps) and outside detected sleep. Outside-sleep intervals do not necessarily represent confirmed wakefulness. Bars show mean ± SEM for 261 matched pairs. Between-group comparisons used paired t-tests with Holm adjustment across five comparisons. ***Adjusted P <0.001; ns, not significant. Glucose axes begin at 100 mg/dL. Blue, lean; orange, obesity.

### Diurnal glucose differences persist with tighter matching on mean glucose

To assess the robustness of the findings to the mean-glucose matching criterion, we repeated one-to-one matching using maximum within-pair differences of 10, 8, 6, 5, 4, and 2 mg/dL, with all other matching criteria unchanged (Fig. 6). The number of matched pairs decreased from 261 at the primary 10 mg/dL threshold to 141 at the strictest 2 mg/dL threshold. Higher nighttime and lower afternoon glucose in participants with obesity persisted across all six thresholds. At the strictest threshold, the nighttime difference was 3.86 mg/dL, compared with 6.18 mg/dL in the primary analysis, while the afternoon difference was −4.27 versus −5.11 mg/dL. The corresponding between-group difference in the night-minus-afternoon contrast decreased from 11.29 to 8.13 mg/dL. All three comparisons remained statistically significant at every threshold after Holm adjustment across the six thresholds separately for each outcome (all adjusted *P* <0.001). Thus, although effect sizes were attenuated under the strictest matching criterion, the opposing nighttime and afternoon glucose differences remained statistically significant.

**Figure 6.**
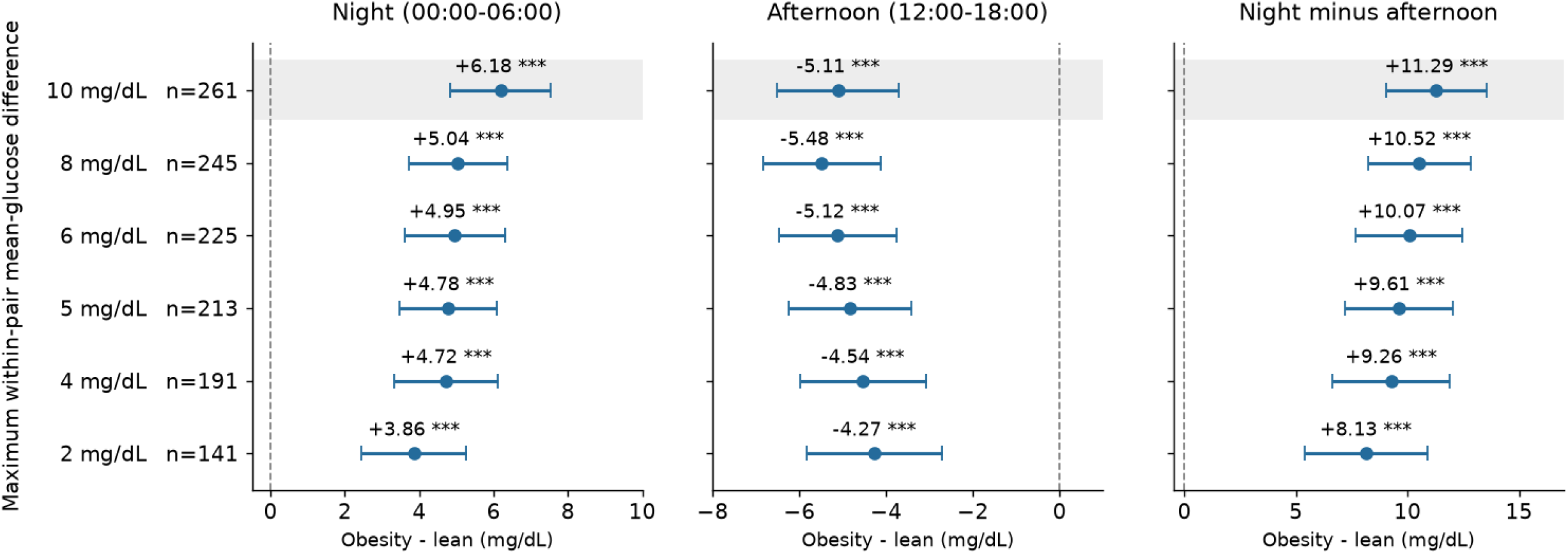
Sensitivity to tighter matching on mean glucose. Paired mean differences (obesity – lean) are shown for nighttime (00:00–06:00), afternoon (12:00–18:00), and the within-participant night-minus-afternoon glucose contrast. One-to-one matching was repeated at each mean-glucose threshold, with all other matching criteria unchanged. Points and error bars represent mean differences and 95% confidence intervals; n indicates matched pairs. Shading identifies the primary analysis (10 mg/dL threshold), and dashed lines indicate zero difference. Comparisons used two-sided paired t-tests with Holm adjustment across six thresholds separately for each outcome. ***Adjusted P <0.001. Samples overlap across thresholds and do not represent independent replications.

## Discussion

Among participants with comparable average glycemia, obesity was associated with higher nighttime but lower afternoon glucose, a pattern we refer to as diurnal glycemic redistribution. This pattern reflects a difference in temporal distribution rather than a uniform elevation. Higher glucose in participants with obesity persisted after alignment to individual sleep onset, and complementary analyses showed higher glucose during wearable-detected sleep but lower glucose outside detected sleep. The opposing nighttime and afternoon differences remained statistically significant with tighter mean-glucose matching, although effect sizes were attenuated. Together, these findings identify an obesity-associated temporal glucose pattern that is not apparent from average glycemic exposure alone.

A key strength was the glycemia-matched design, which enabled comparison of temporal glucose patterns at comparable average exposure. The cohort comprised 261 matched pairs (522 participants) across the healthy, prediabetes/lifestyle-controlled, and non-insulin-medication categories, allowing examination within distinct clinical groups rather than only among participants receiving diabetes treatment. Repeated free-living CGM measurements characterized glucose profiles across multiple days, while linked wearable records provided an individualized reference for sleep timing. This integration allowed glucose differences to be assessed relative to sleep onset and during detected sleep, without assuming that fixed nighttime intervals represented sleep. Concordant findings across these complementary temporal definitions, together with persistence under tighter mean-glucose matching, support the consistency of the observed pattern within this cohort.

The physiological and behavioral determinants of diurnal glycemic redistribution remain unresolved. Differences in glucose production and disposal, meal timing, and sleep-related factors are possible contributors, but their roles cannot be distinguished by the present analyses. Nevertheless, comparable average glycemia need not imply comparable temporal glucose regulation. The pattern observed within the dataset-defined healthy category raises the possibility that temporal glucose features could complement conventional measures in identifying otherwise unrecognized metabolic vulnerability. Such differences do not, by themselves, establish dysfunction or increased disease risk. Longitudinal studies are needed to determine whether diurnal glycemic redistribution precedes glycemic deterioration and provides predictive information beyond HbA1c, mean glucose, and established CGM metrics.

Several limitations should be considered. First, the cross-sectional observational design does not establish causality or determine the physiological mechanisms underlying diurnal glycemic redistribution. Second, matching on mean glucose improves comparability but may introduce selection and does not eliminate residual confounding from factors such as meal timing, physical activity, sleep behavior, or medication use. Because average glycemia was constrained by design, higher glucose during one part of the day must be offset to some extent during another; however, the specific timing and magnitude of the nighttime and afternoon differences were not imposed by the matching procedure. Third, wearable-derived sleep measures may not fully capture physiological sleep or circadian phase. Finally, the findings were derived from a single cohort and require validation in independent populations. Despite these limitations, the results show that comparable average glycemia can conceal systematic differences in the temporal distribution of glucose associated with obesity.

In summary, obesity was associated with diurnal glycemic redistribution despite comparable average glycemia, characterized by higher nighttime and lower afternoon glucose. This pattern was evident across clock-time and sleep-related analyses and persisted with tighter glycemic matching. These findings suggest that average glycemic measures can obscure meaningful temporal differences in glucose regulation and support further evaluation of diurnal glycemic redistribution as a potential marker of metabolic vulnerability.

## Statements and Declarations

### Ethical approval

The present study was approved by the Institutional Review Board of Wayne State University.

### Consent to Participate

Not applicable.

### Author Contributions

R.Z. conceptualized, designed the study, analyzed the data, and wrote the paper.

### Ethical Responsibilities of Authors

The author has read, understood, and complied, as applicable, with the statement on Ethical Responsibilities of Authors in the Instructions for Authors.

### Data Availability Statement

The data analyzed in this study were obtained from AI-READI version 3.0.0, available through FAIRhub at https://doi.org/10.60775/fairhub.3. Access and reuse are subject to the dataset license and applicable access requirements.

